# Safety and efficacy of frontline doxorubicin in combination with temozolomide and valproic acid for the treatment of pediatric malignant gliomas: results of a phase 2 study

**DOI:** 10.1101/2025.09.15.25335744

**Authors:** B. Castelli, M. Tellini, M.A. Malanima, M. Guidi, L. Giunti, C. Fonte, M. Di Nicola, M.L. Censullo, F. Giordano, I. Desideri, D. Greto, S. Ricci, L. D’Incerti, C.G. Gori, A. Pugi, K. Tortora, B. Tirinnanzi, C.E. La Torre, E. Pasquinelli, R. Amato, M. Scagnet, L. Genitori, A. Iacono, A.M. Buccoliero, E. Bennati, S. De Masi, I. Sardi

## Abstract

**Background:** Despite innovative approaches, outcomes for pediatric high-grade gliomas (HGGs) remain poor. Doxorubicin (Dox) is commonly used to treat many childhood cancers, with a well-known safety profile, although the blood-brain barrier limits its use in central nervous system tumors. However, its antineoplastic activity is reported *in vitro* and *in vivo* glioma models. We aimed to assess safety and activity of the addition of Dox to the standard treatment in this population.

**Methods:** A monocentric, non-randomized, phase II interventional study was opened at Meyer Children’s Hospital IRCCS in Florence (EudraCT 2015-002307-28), introducing Dox 100 mg/m^2^ over a 96-hour infusion following chemo-radiotherapy as a post-operative treatment, alongside valproic acid throughout the treatment. The endpoints were safety and efficacy of the add-on Dox approach in prolonged infusion.

**Results:** Twenty-one heterogeneous malignant pediatric HGGs patients were enrolled. However at the time of Dox administration, only twelve patients presented a performance status sufficient to receive the investigational drug. Dox single course-group (10 patients) exhibited a median overall survival (OS) of 13.7 months (6.9 months in non-Dox-treated patients). Analyzing a multivariate Cox regression, patients with diffuse midline glioma showed a significantly higher risk of events compared to those with other HGG (approximately 80%, p = 0.008). Dox-treated DMG shows a slight reduction in event rate (9.52 vs 12.55). Interestingly, all patients (6/12) with hemispheric malignant glioma, who had undergone Dox, relapsed at sites distant from the primary tumor. Currently, only one patient is alive (a Dox-treated grade 3 anaplastic pleomorphic xanthoastrocytoma), Considering the Dox-treated patients, despite 35 Serious Adverse Reactions related to Dox were reported, predominantly hematologic, the treatment after focal radiotherapy was well tolerated. No signs of cardiotoxicity, nephrotoxicity, or neurotoxicity following Dox infusion were reported.

**Conclusion:** This preliminary study shows that a prolonged infusion Dox add-on to standard multimodal treatment for pediatric HGGs is well tolerated with no significant adverse events and with a positive impact in terms of survival, although not statistically significant.

## Introduction

Pediatric high-grade gliomas (HGGs) are heterogeneous and highly aggressive brain tumors, representing one of the most lethal malignancy in pediatric oncology [1,2]. They make up 15–20% of pediatric central nervous system (CNS) tumors [2]. Despite intense research, prognosis remains extremely poor, with a five-year survival rate of less than 20% [3,4]. The consolidated first-line treatment for pediatric and young adult HGGs typically involves a multimodal approach aimed at maximizing patient outcomes. It includes maximal safe surgical resection if feasible, followed by focal radiotherapy (RT) approach associated with concomitant and adjuvant temozolomide (TMZ) according to Weller-Stupp strategy [5]. Location, aggressiveness, and diffuse infiltrative growth make glioblastoma (GBM) therapy extremely challenging [4]. Given these critical features, there is a compelling need for innovative and effective therapeutic strategies [4]. Indeed, despite advances in the treatment of GBM, outcomes remain unsatisfactory, and the current standard of care continues to heavily rely on chemotherapy following optimal surgical removal [6].

Doxorubicin (Dox) is a well-established antineoplastic agent used in many tumors, together with other chemotherapy agents [4]. Dox is an anthracycline antibiotic, with anticancer activity related to topoisomerase II inhibition, through intercalation into double-strand DNA [7]. Its clinical wide application is limited due to severe side effects, mediated by oxidative stress, apoptosis and inflammation [8,9]. The heart is a preferential target of Dox toxicity, such as heart failure, arrhythmia, and myocardial infarction [8,10]. Oxidative stress, altered autophagy, inflammation, and apoptosis/ferroptosis represent the main pathogenetic mechanisms responsible for Dox-induced cardiotoxicity (DIC) [8,10]. Apart from cardiotoxicity, Dox induces vascular toxicity, represented by arterial stiffness and endothelial dysfunction [11]. This anticancer drug also affects other organs including the brain, kidney and liver [12]. It has been reported that Dox may cause renal failure (Dox-induced nephrotoxicity) and there is no effective treatment currently available for Dox-induced kidney damage [13]. Acute Dox exposure dose-dependently impairs synaptic processes associated with hippocampal neurotransmission, induces apoptosis, and increases lipid peroxidation resulting in neurotoxicity [14]. The occurrence of chemo brain and impairment of neurocognitive functions have also been debated, via indirect Dox-mediated neurotoxicity mechanism [15,16]. Reports of posterior reversible encephalopathy syndrome (PRES) associated with the use of anthracycline containing chemotherapeutic agents have been published, in particular following DA-EPOCH (dose-adjusted etoposide, prednisone, vincristine, cyclophosphamide, doxorubicin) or ABVD (doxorubicin-adriamycin, bleomycin, vinblastine and dacarbazine) [17-19]. Different strategies have been proposed to attenuate toxicity, including combined therapy with bioactive compounds [20].

CNS tumors seem to be resistant to Dox use [21]. The poor outcomes in CNS tumors should be in part explained by the ineffective delivery of anticancer agents, as Dox, into the CNS across the blood brain barrier (BBB) [4]. The ATP-binding cassette (ABC) transporters-mediated active efflux of anticancer drugs may form a first line of pediatric HGGs treatment resistance [2,4]. *In vitro* data demonstrate that Dox is toxic to GBM cell lines and *in vivo* data in animal models confirm that locally delivered Dox is effective as monotherapy against experimental intracranial glioma [22]. Given the sensitivity of gliomas to Dox, it is crucial to address the mechanisms of resistance that can limit its efficacy. Struggles to develop modalities to expose the entire tumor to pharmacologically meaningful quantities of therapeutics have been attempted. Prolonged exposure to anthracyclines (96-hours) seems to induce a significant apoptosis rate in resistant GBM stem cells [21]. In glioma progenitors, after prolonged drug exposure (96h), Eramo et al. observed a significant induction of apoptosis after anthracyclines compared to commonly used etoposide and TMZ [23]. In addition, RT can cause BBB disruption, resulting in an increased permeability into the surrounding brain parenchyma [24,25]. In the context of brain cancer, an increased BBB permeability should indeed offer therapeutic advantages, permitting otherwise BBB-blocked drugs to better reach glioma cells [26]. Both clinical and preclinical studies revealed a peak in permeability due to vascular damage a few months after radiation, followed by BBB restoration afterwards [26-28]. Farjam et al (2015) and Cao et al (2009) noticed a peak in permeability at 1–1.5 months, which was reversed over time [26,28,29]. Yuan et al. (2006) demonstrated that BBB permeability was significantly increased 90 days after fractionated radiotherapy (FRT) and continues to rise until 180 days post-treatment [24]. Intriguingly, preclinical studies also reported that valproic acid (VPA) may affect tumor cells, inhibiting proliferation, and angiogenesis, and promoting apoptosis and autophagy [30]. VPA could exert an antineoplastic action mainly as a histone deacetylase (HDAC) inhibitor, resulting in chromatin remodeling, but it also displays DNA-demethylation properties [31]. It enhances TMZ antitumor effect in TMZ-resistant malignant glioma cells, through a possible VPA-mediated downregulation of MGMT expression [31,32]. The combination of VPA, TMZ and RT has been reported to cause a significant radiation enhancement, without antagonizing the cytotoxic effects of TMZ [31,33]. In 2020 in a phase 2 study Su et al. evaluated valproic acid in association with RT in children diagnosed with HGGs [34]. Based on these considerations, we aimed to introduce Dox add-on therapy with concomitant VPA to standard treatment in pediatric HGGs, investigating whether 96h continuous Dox infusion displays a low-acceptable toxicity and evaluating the efficacy of the combined regimen.

### Patients and methods

This was an open-label monocentric not-randomized phase II interventional study (GBM TMZ/DOX 2015 - EudraCT 2015-002307-28). The study was approved by “Comitato Etico Regione Toscana – Pediatrico”, the ethics committees of the participating center. All patients provided written informed consent. The study was supported by the grant of Ministry of Health “All ages malignant glioma: holistic management in the personalized minimally-invasive medicine era – from lab to rehab (GLI-HOPE - NET-2019-12371188)”. The endpoints were to evaluate the tolerability and efficacy of prolonged Dox infusion in conjunction with standard Weller-Stupp treatment. All data reported to the Institute of Pharmacovigilance were analyzed.

### Eligibility

From February 2016 to January 2020 all patients aged 3-30 years with histological documented diagnosis of high-grade glioma (HGGs) according to WHO 2016-2021 classification [35] were eligible for the study, provided they satisfied the following criteria: Karnofsky/Lansky performance status of 40% or greater, and adequate hematologic, renal, and hepatic function (absolute neutrophil count ≥1500/mm^3^; platelet count ≥100.000/mm^3^; serum creatinine level ≤1.5 × ULN; total serum bilirubin level ≤1.5 × ULN; and liver-function values <3 × ULN), normal electrocardiogram and echocardiography (ejection fraction >60 % and shortening fraction >30 %), and no organ toxicity with WHO grade >2. No other anticancer treatment was allowed during the study. Notably, diagnosis of diffuse intrinsic pontine glioma (DIPG) was based primarily on magnetic resonance imaging in most cases [36]. Previous other treatments were not allowed.

### Treatment schedule

At diagnosis all patients were treated in accordance with Weller-Stupp protocol [5]. RT involved focal normo-fractionated irradiation given once daily, five days per week over a period of six weeks, to a total dose of 54-59.4 Gy. Concomitant chemotherapy consisted of TMZ at a dose of 75 mg/m^2^/day, administered daily for 7 days a week, from the first to the last day of radiotherapy.

Following a 4-week break from the completion of the radiotherapy/concomitant TMZ phase, in the first version of the protocol, starting from week 10, patients received Dox given over a 96-hour continuous infusion at the dose 25 mg/m^2^/day (cumulative dose 100 mg/ m^2^/cycle) for 3 cycles every 28 days. A subsequent protocol amendment (March 2, 2017) introduced an adjuvant TMZ cycle at week 10, prior to Dox administration and reduced the total number of adjuvant Dox cycles to one, due to a prolonged although reversible grade 4 multilineage cytopenia. After four weeks from the completion of Dox treatment, adjuvant TMZ treatment was resumed according to standard of care protocol for 16 post-Dox cycles every 28 days. During the entire treatment course, patients were taking valproic acid 20-30 mg/kg/day divided in two oral doses (Figure 1).

**Figure 1.**
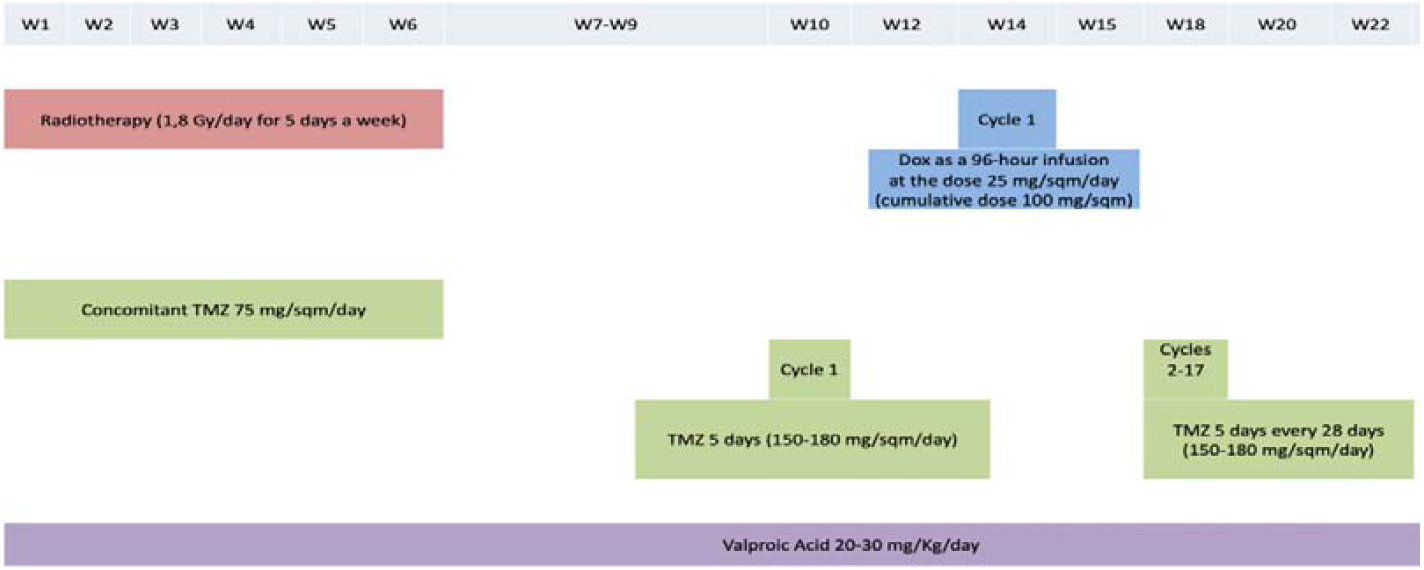
Treatment schedule GBM TMZ/DOX 2015 (version March 2, 2017).

### Toxicity

Toxicities were measured according to Common Terminology Criteria for Adverse Events v4.03 (CTCAE). Clinical examination, full blood count, liver, renal, and electrolyte values, was performed before and after each course. Hematological toxicity was assessed until normalization of full blood counts. Cardiological consultation was performed at baseline and after the completion of Dox treatment or previously in case of necessity, apart from patients who have suffered from a decline due to a rapid disease progression.

### Tumor response

Response was assessed by brain contrast-enhanced MRI performed every three months during treatment or in relation to clinical status and it was formulated according to RAPNO criteria(37). Objective responses were to be confirmed at 4–6 weeks. Efficacy parameters included complete response (CR), partial response (PR), duration of response (time from CR or PR to disease progression), progression-free survival (PFS) and overall survival (time from first treatment to disease progression or to death/cutoff date, respectively).

### Statistical analysis

PFS and OS rates were calculated using the Kaplan–Meier method. Subgroup analysis was performed, to evaluate the effects of the treatment within specific subsets of the study population. In univariate analysis using the Cox regression model, we assessed the relationship between each variable of interest and time-to-event (progression or death). We presented the results of a multivariate Cox regression analysis both with and without interaction terms.

## Results

### Patient characteristics

Between October 2015 and January 2021, 21 patients (twelve males, nine females) were enrolled, with a median age of 7 years (range 3-22 years). The main clinical and histopathological features of this cohort are summarized in Table 1. The tumor location was cerebral hemisphere in six patients, thalamic in two, brainstem in nine. Four patients presented with gliomatosis cerebri. One patient had spinal metastatic disease at diagnosis, three patients presented leptomeningeal enhancement. Twelve diagnoses were histological: two anaplastic astrocytomas (one IDH1 R132H mutant), three high grade gliomas H3G34 mutant, four high grade gliomas H3-wildtype IDH-wildtype, one grade 3 anaplastic pleomorphic xanthoastrocytoma, two diffuse midline gliomas (DMG) H3K27M-mutant. Regarding the extension of the surgical approach prior to inclusion in the study, five patients were subjected to biopsy, six patients to partial resection and one patient to gross tumor removal. Four out of 21 tumors (19%) have arisen in the context of cancer predisposition syndromes: two Li Fraumeni syndrome, one RASopathy, one Ollier disease.

**Table 1.** Main demographic and histological features of this series.

| Patient | Age range at enrollment (years) | Sex | Cancer Histology | Site | Dox YES/NO n. of cycles |
| --- | --- | --- | --- | --- | --- |
| 1 | 6-10 | M | NA | Midline | NO |
| 2 | 6-10 | M | NA | Midline | YES (III) |
| 3 | 6-10 | F | NA | Midline | YES (III) |
| 4 | 11-15 | M | HGG H3 G34 mutant | GC | NO |
| 5 | 1-5 | M | NA | Midline | NO |
| 6 | 6-10 | F | NA | Midline | YES (I) |
| 7 | 16-20 | F | Anaplastic Astrocytoma IDH mutant | GC | NO |
| 8 | 6-10 | F | DMG H3 K27-altered | Midline | NO |
| 9 | 1-5 | F | NA | Midline | NO |
| 10 | 6-10 | M | NA | Midline | NO |
| 11 | 11-15 | F | HGG H3 G34 mutant | GC | YES (I) |
| 12 | 11-15 | M | HGG H3 G34 mutant | GC | YES (I) |
| 13 | 11-15 | M | HGG H3-wildtype IDH-wildtype | GC | NO |
| 14 | 11-15 | M | HGG H3-wildtype IDH-wildtype | Hemispheric | YES (I) |
| 15 | 6-10 | F | DMG H3 K27-altered | Midline | YES (I) |
| 16 | 6-10 | M | HGG H3-wildtype IDH-wildtype | Hemispheric | YES (I) |
| 17 | 6-10 | M | HGG H3-wildtype IDH-wildtype | GC | YES (I) |
| 18 | 1-5 | M | NA | Midline | YES (I) |
| 19 | 6-10 | F | Anaplastic pleomorphic xanthoastrocytoma | Hemispheric | YES (I) |
| 20 | 6-10 | M | Anaplastic Astrocytoma | GC | YES (I) |
| 21 | 6-10 | F | NA | Midline | NO |

M: male; F: female; NA: not applicable; HGG: high-grade glioma; DMG: diffuse midline glioma; GC: gliomatosis cerebri

### Study treatment and safety

Considering the 21 enrolled patients, only 12 (57.14%) were able to undergo experimental treatment with Dox. Nine did not have a satisfactory performance status (PS) at the time of Dox administration or experienced early disease progression. Two patients (2/21) received three Dox courses according to the first protocol and ten patients (10/21) received a single course in accordance with the revision of the protocol, so that 16 courses were administered in the 12 patients. The protocol amendment was necessary due to prolonged, albeit reversible, hematological toxicity.

Safety analysis included all the subjects of the study (n = 21): 65 Serious Adverse Event (SAE) were reported. Nineteen of 21 patients (19/21) presented at least one SAE.

Considering the 12 Dox-treated patients, each patient developed at least one SAE, for a total of 52 SAEs. Moderate and severe SAEs were predominantly hematologic. Six out of twelve Dox-treated patients (6/12) experienced at least one serious adverse event considered related to Dox (Serious Adverse Reaction, SAR). In total SARs were 35. Twenty out of thirty-five SARs (20/35) were reported in the two patients treated with three Dox courses according to first protocol version (all G3 and G4 toxicities). The long lasting multilineage cytopenia was reversible but it exposed patients to a higher risk of febrile neutropenia, mucositis, transfusion support and hospitalization in general, which lead us to revise the study design.

There were not Suspected Unexpected Serious Adverse Reactions (SUSAR). All patients discontinued the study due to disease progression. No anticipated Dox suspension was reported. Two patients interrupted concomitant TMZ for hematological toxicity.

Post-treatment echocardiographies were performed after Dox cycle in 6 patients and were always within normal ranges. No patients showed nephrotoxicity, hepatotoxicity or neurotoxicity following Dox infusion. The occurred neurologic deteriorations were in the context of disease progression rather than secondary to study treatment.

### Study treatment and efficacy

Considering the entire population, the median OS, generated using the Kaplan-Meier method, was 9.7 months (Figure 2) and after a follow up of 75.4 months, 20 out of 21 patients had died (stratified analyses according to Dox administration and tumor type are reported in Supplementary Tables 1-2). Dox single course-group exhibited a median OS of 13.7 months, conversely patients who did not receive Dox demonstrated a median OS of 6.9 months, however this difference was not statistically significant, since eligibility to Dox treatment was mainly hinder by a more aggressive tumor behavior after radiotherapy (Table 2). Additionally, there was no significant difference when comparing patients treated with a single course of Dox to those who were not treated, stratified by tumor histology dividing in diffuse midline glioma (DMG) vs. other types of HGG, sex, or age at onset (Table 2).

**Figure 2.**
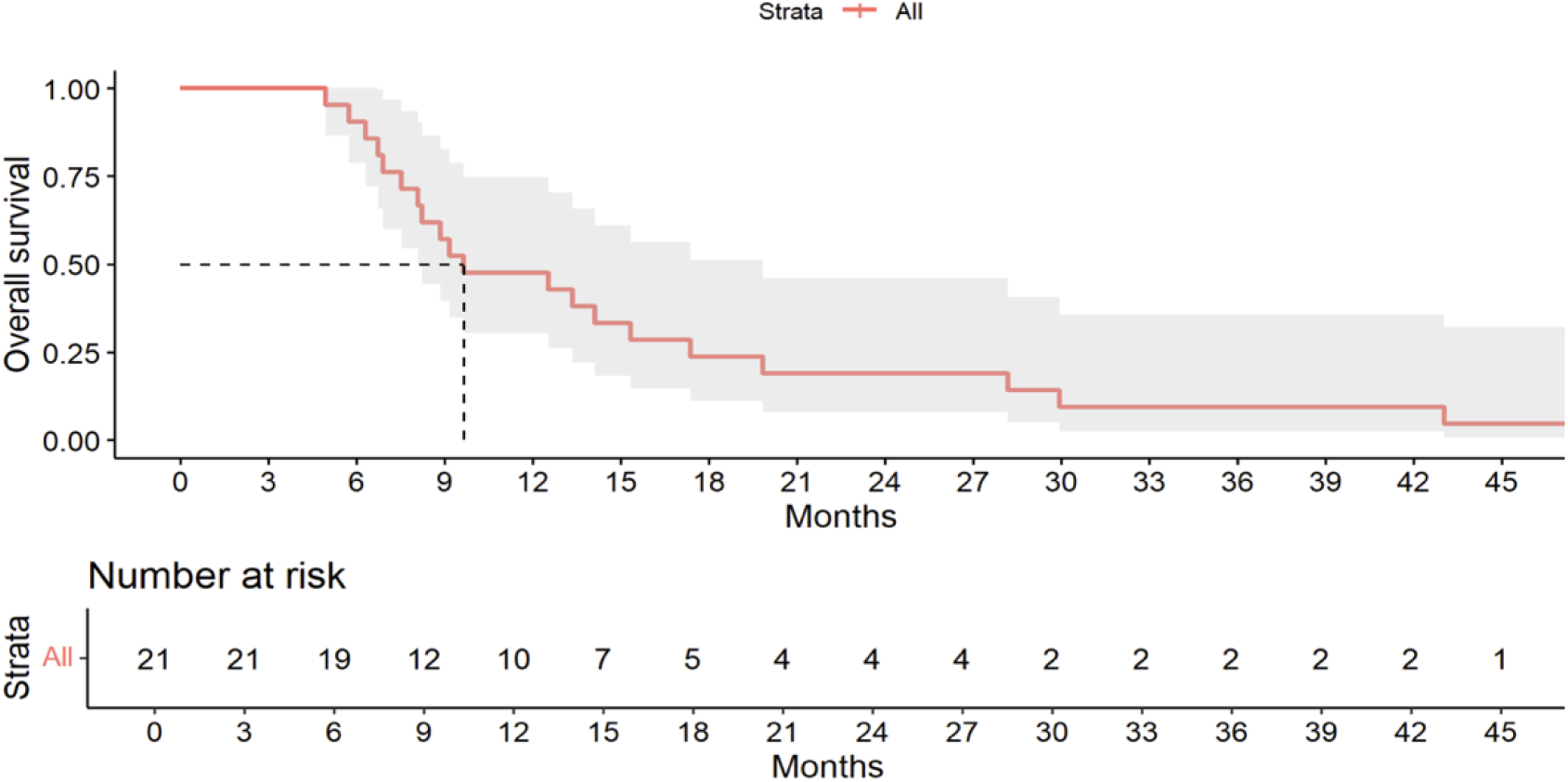
Kaplan-Meier survival curve for all 21 patients. The median overall survival was 9.7 months (95% CI: 8.1– 19.8), with 21 patients and 20 events. The table below the curve shows the number of patients at risk at each time point.

**Figure 3.**
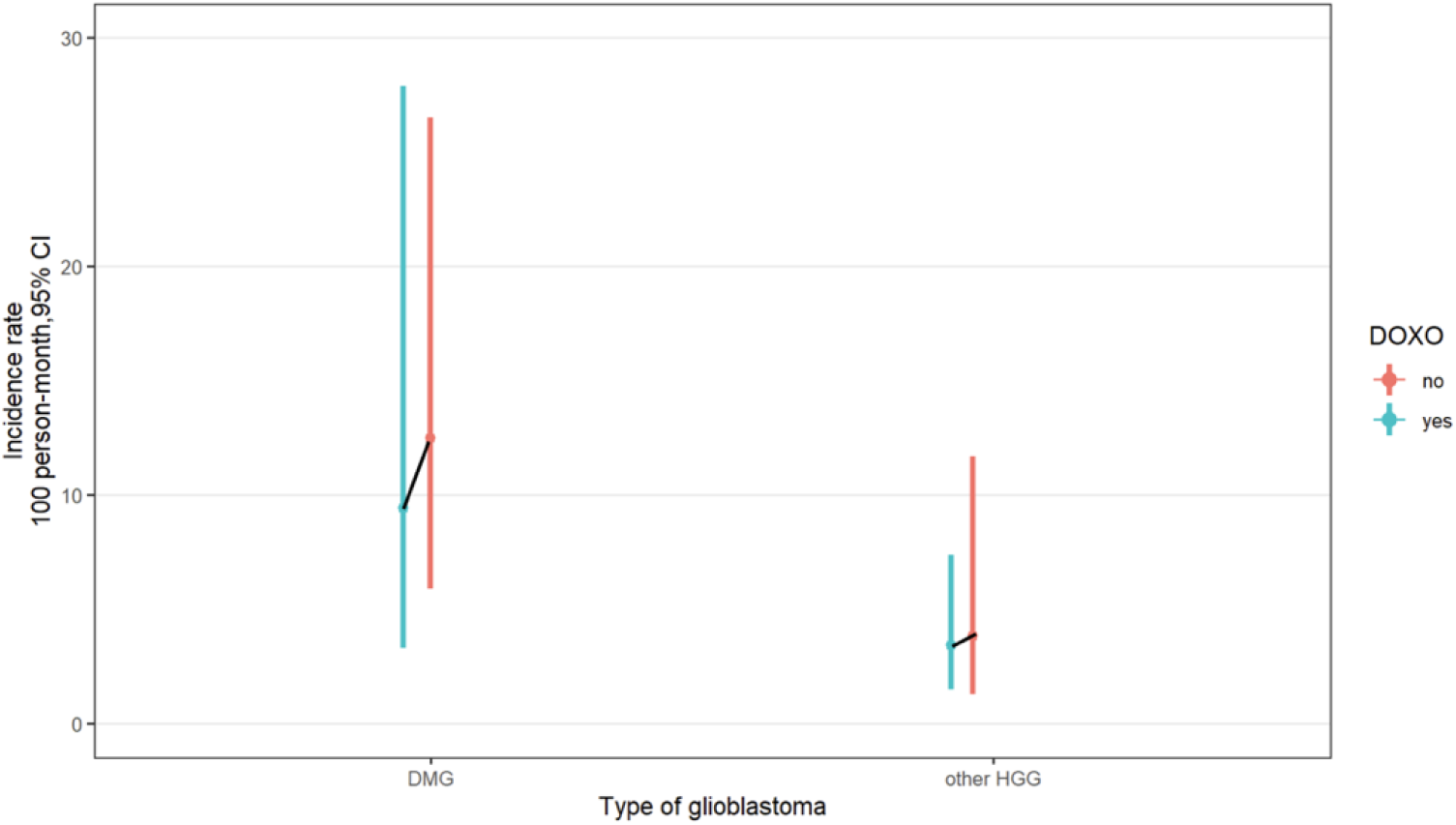
Incidence rates in Dox and no Dox groups stratified by type of HGG.

**Table 2.** General Characteristics of 21 Patients Stratified by Doxorubicin Administration.

|  | No Dox n=9 | Dox (1 dose) n=10 | p-value* |
| --- | --- | --- | --- |
| Type of HGG |  |  |  |
| DMG | 6 (66.7) | 3 (30.0) | 0.18 |
| OtherHGG | 3 (33.3) | 7 (70.0) |  |
| Sex |  |  |  |
| Female | 4 (44.4) | 4 (40.0) | 1 |
| Male | 5 (55.6) | 6 (60.0) |  |
| Onset (years) |  |  |  |
| < 12 | 6 (66.7) | 6 (60.0) | 1 |
| ≥12 | 3 (33.3) | 4 (40.0) |  |
| Event |  |  |  |
| Yes | 9 (100) | 9 (90.0) | 1 |
| No | 0 (0) | 1 (10.0) |  |
| Survival (median months [IQR]) | 6.9<br>[6.3, 15.3] | 13.7<br>[10.0, 19.2] |  |

Note: Dox: doxorubicin, HGG: high-grade glioma, DMG: diffuse midline glioma.
\* The p-values refer to the statistical tests comparing variables between patients who did not receive Doxorubicin (No Dox) and those who received one dose of Dox.
The fourth column presents the p-values, indicating the statistical significance of differences in the variables of interest between patients who received one dose of Dox and those who received none.

Table 3 displays the event rates (per 100 patients) of all 21 patients, as well as for groups defined by Dox administration. Additionally, for those who did not receive the drug or received a dose (19 patients), stratified rates were calculated based on categories of interest, including cancer type, age at onset, and sex. Dox-untreated DMG represents the highest risk of the event. Dox-treated DMGs show a slight reduction in event rate (9.52 vs 12.55) (Table 3). In our study, the cohort under 12 years was found to be correlated with a higher probability of the event, but only in untreated patients (Table 3)(the cumulative survival probabilities stratified by Dox treatment are reported in Supplementary Table 4).

**Table 3.**
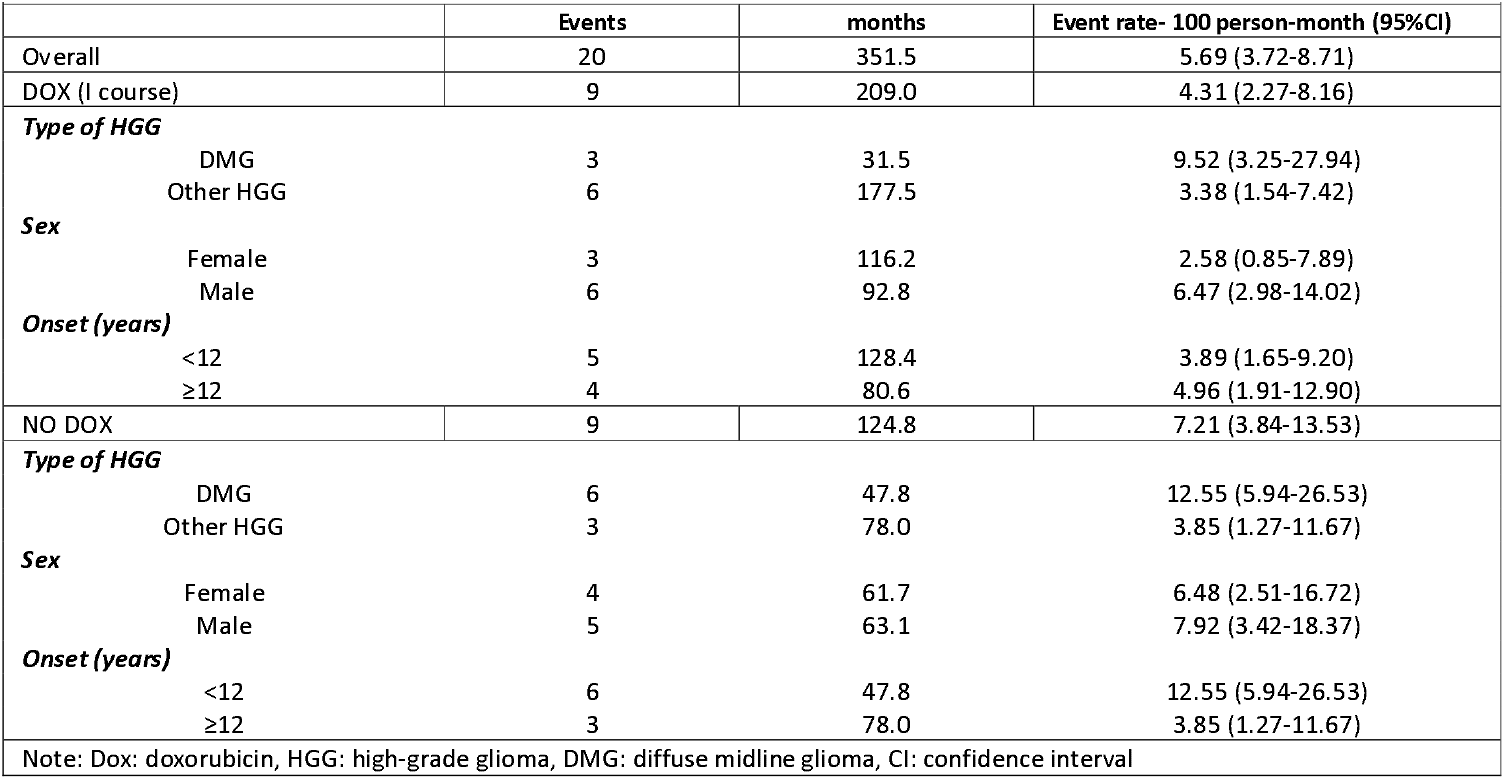
Overall event rate and stratified event rate by Dox administration with 95% CI.

The risk of events was analyzed using a univariate Cox regression model, focusing on several variables of interest, including drug administration, type of HGG, age at onset, and gender (Table 4). The results indicate that patients with DMG have a significantly higher risk of events compared to those with other HGG (about 80% less, p = 0.008). These data are consistent with findings reported in the literature [38]. Dox-treated group showed a slight reduction of the hazard ratio; however statistical significance was not reached (Table 4).

**Table 4.** Univariate Cox regression analysis for characteristics of interest

|  |  |  |  |
| --- | --- | --- | --- |
| **Variable** | **Category** | **HR (95% CI)** | **p-value** |
| Doxorubicin | No Dox | 1 (ref) | 0.24 |
|  | Dox | 0.57 (0.22–1.45) |  |
| Type of HGG | DMG | 1 (ref) | 0.008 |
|  | Other HGG | 0.19 (0.01–0.65) |  |
| Sex | Female | 1 (ref) | 0.46 |
|  | Male | 1.46 (0.54–3.99) |  |
| Onset (years) | <12 | 1 (ref) | 0.14 |
|  | ≥12 | 0.48 (0.17–1.29) |  |
| Note: Dox = doxorubicin; HGG = high-grade glioma; DMG = diffuse midline glioma; HR = hazard ratio; CI = confidence interval. |  |  |  |

Table 5 shows the results of the multivariate model, which includes both the Dox variable and the type of HGG (DMG vs other HGG), along with the interaction term between these two variables (the model without interaction term is reported in Supplementary Table 3). After verifying the proportional hazards assumptions for the variables, we report the coefficients and their corresponding confidence intervals from the Cox model. The reference category used to calculate the risk was Other HGGs Dox treated. In no Dox treated Other HGGs, the risk resulted very similar to those who receive Dox and had other HGG (HR=0.94, p=0.93). In DMGs who received Dox the risk was higher, but not significantly (HR=3.43, p=0.12). In DMGs who do not receive Dox, the risk was significantly higher (HR=6.65, p=0.006). Interestingly, all patients (6/12) with hemispheric malignant glioma, who had undergone Dox, relapsed distant from the primary tumor. To date, after 75.4 months of follow up, only one Dox-treated patient with anaplastic pleomorphic xanthoastrocytoma is alive.

**Table 5.**
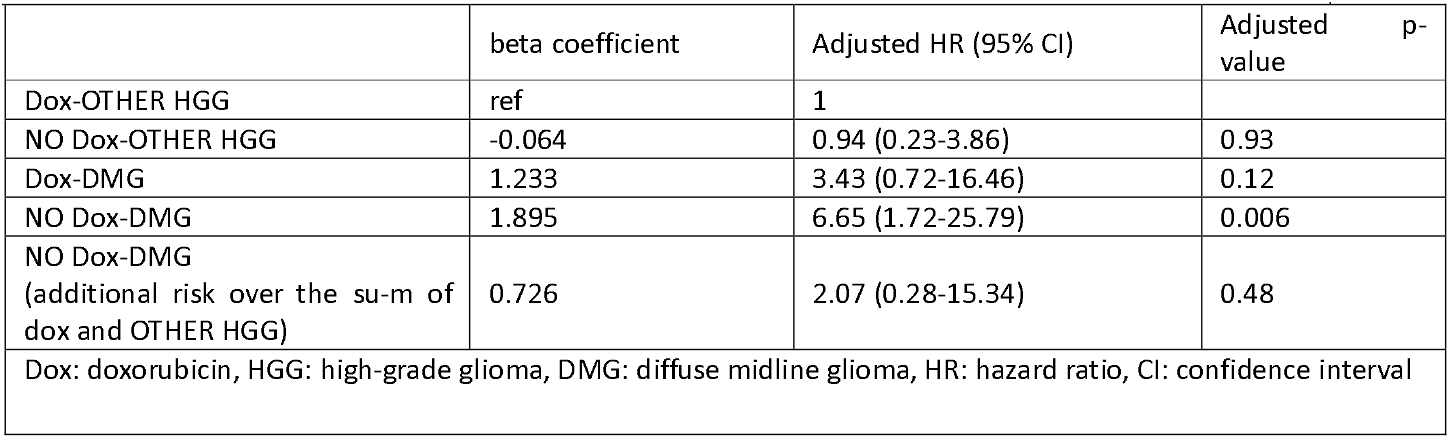
Results of multivariate Cox regression for the characteristics of interest with the interaction term.

## Discussion

Pediatric high-grade gliomas (HGG), including DMGs, are one of the leading cause of cancer-related death in children [2]. Several drugs have shown great toxicity in glioblastoma (GBM) *in vitro* models, but clinical trials, exploring consolidated and innovative agents, have shown no significant improvement in terms of OS [4].

Anthracyclines represent the class of antitumor drugs with the broadest spectrum of activity in pediatric oncology [7]. Given the deep experience in clinical practice, major concerns regarding Dox use in malignancies with dismal prognosis are related to management of acute toxicities with morbidity and hospitalization risk and long-term dose-dependent cardiotoxicity and neurotoxicity [17,39,40].

The “chemo-brain” effect sheds light on Dox-based interference with neural function of a drug that notoriously shows a certain degree of inability to cross the blood-brain-barrier (BBB) [21]. On the other hand, there is a strong preclinical background for Dox effectiveness in malignant glioma provided by different groups. Veringa et al. demonstrated that Dox shows a high cytotoxic effect on primary GBM cell lines, more than conventional used TMZ and etoposide [4,2]. Preclinically, Aldoxorubicin (AlDox), a novel Doxorubicin prodrug, has been successfully tested against GBM, encouraging the study of its association with other agents [41]. In recent years, our group has also contributed to the investigation on Dox efficacy in the treatment against GBM, reporting how Dox, in combination with other drugs, has a high cytotoxic effect against GBM cell lines and orthotopic xenograft mice model of human GBM [4]. We investigated the combination of Dox and rapamycin (Rapa), *in vitro* and *in vivo* GBM models, testing the advantage of the addition of the antiproliferative/cytotoxic effect of Rapa with the cytotoxic effect of Dox [4]. Oraiopoulou et al. suggested a TMZ–Dox dual chemotherapeutic scheme to both halt proliferation and increase cytotoxicity against GBM, using patient□derived spheroids [42]. In 1999 Stan et al. provided evidence that two highly invasive human glioma cell lines (U-87 MG and U-373 MG) enter apoptosis after 48 hours following 24 h growth arrest induced by Dox [43]. In 2005 Lesniak et al. demonstrated that Dox, when locally delivered, is an effective monotherapeutic agent against experimental intracranial glioma [22].

In addition to inability to cross the BBB, however, drug efflux transporters at the BBB may represent a key mechanism of resistance of pediatric HGGs to such treatment [2]. In 2013 Veringa et al. explored the pathway of Dox drug resistance, focusing on the hampering presence of P-glycoprotein (P-gp, ABCB1), breast-cancer-resistance protein (BCRP, ABCG2), and multidrug-resistance-associated proteins (MRPs, ABCC1) [2]. Considering that, in 2016 our group evaluated the antitumor efficacy of a combination treatment of Dox and morphine using an orthotopic GBM xenograft model, proceeding from the fact that morphine seems to mediate the uptake of Dox into the brain by its reduced efflux mediated by P-gp and inducing a transient alteration of the permeability of BBB [21]. Regarding other bypassing mechanisms Liao et al. used the shockwave system to enhance the BBB-crossing capacity of Dox and investigated its therapeutic effects on rats with GBM [44]. The characteristics of Dox release from polymeric carriers was also investigated [45]. Various strategies have been emerged to enhance the therapeutic use of the drug, including the incorporation in nanoscale carriers as liposomes, polymer and peptide/protein conjugates, polymeric micelles, nanoparticles [41].

In this study, we aimed to introduce Dox as add-on therapy to standard treatment in pediatric and young adult HGGs, exploiting the toxicity and the pro-apoptotic activity of prolonged infusion of Dox during the frameshift of the peak of radiation-induced BBB opening. Due to study exit or ineligibility at the time of Dox treatment, only twelve out of twenty-one patients received the interventional treatment. Considering the entire population, the median OS was 9.7 months. Dox single course-group exhibited a median OS of 13.7 months, conversely patients who did not receive Dox demonstrated a median OS of 6.9 months, anyway this difference was not statistically significant. The extension of surgical debulking can indeed introduce bias and it can influence prognosis, as the diverse histology. The only patient who survived underwent gross total removal for a grade 3 pleomorphic xanthoastrocytoma.

However, concerning drug safety, a single prolonged course of Dox resulted in tolerable toxicity. This study demonstrates that Dox should exert some effect on malignant gliomas, however, the impact on clinical outcomes needs further analysis due to the great heterogeneity of our cohort.

## Conclusion

*In vitro* and *in vivo* studies showed that HGG, as several other tumors, are expected to be sensitive to anthracyclines. One major obstacle to the use of Dox may involve inadequate drug delivery to the tumor site because of the BBB. Our investigational phase II study, designed on the RTP induced temporary BBB opening, shows that prolonged infusion of Dox in addition to the standard treatment in pediatric and young adult HGGs is feasible and tolerated. These results may open the way to novel therapeutic approaches to refractory gliomas of adult and pediatric age.

## Acknowledgments

We are very grateful to “Festa di Sole” and “La Forza di Gio’” odv for their competent assistance in supporting young researchers.

## Data availability

The research data for this study can be shared upon request from the corresponding author by any qualified investigator.

