## Supplementary_Table 1-2-3-4 for "Safety and efficacy of frontline doxorubicin in combination with temozolomide and valproic acid for the treatment of pediatric malignant gliomas: results of a phase 2 study"

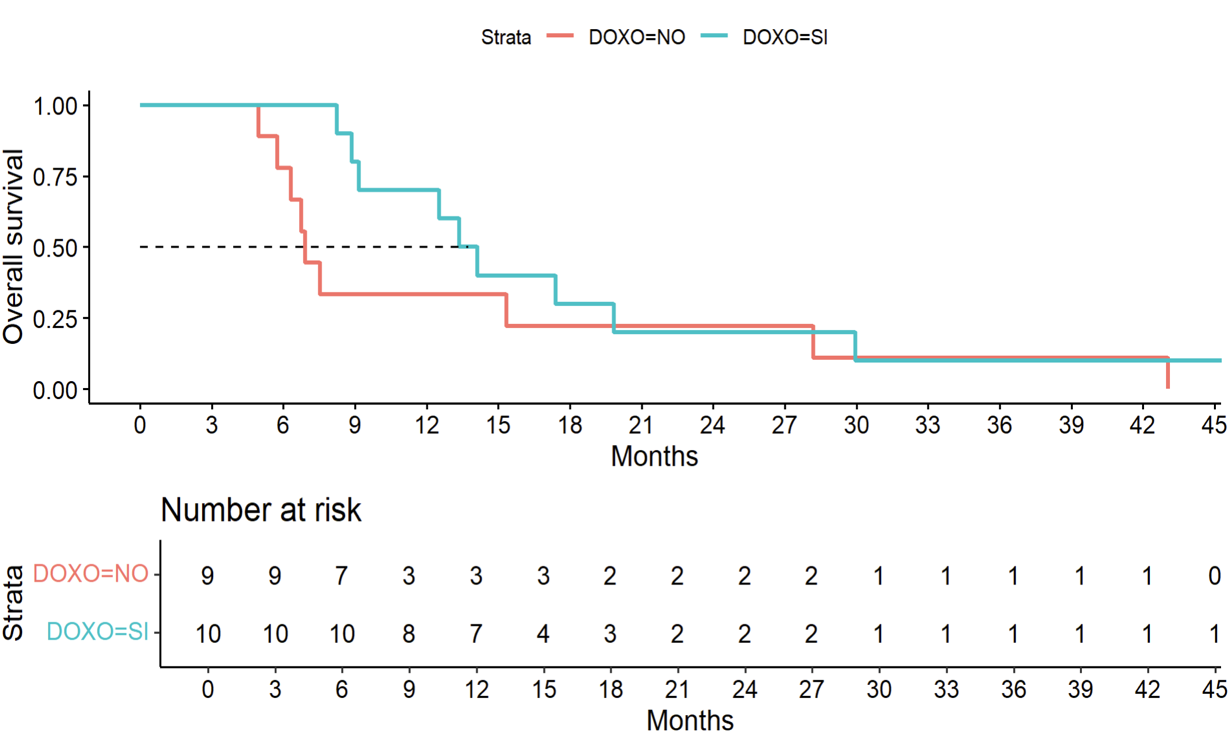

Supplementary material 1. Survival curves stratified by Dox administration.

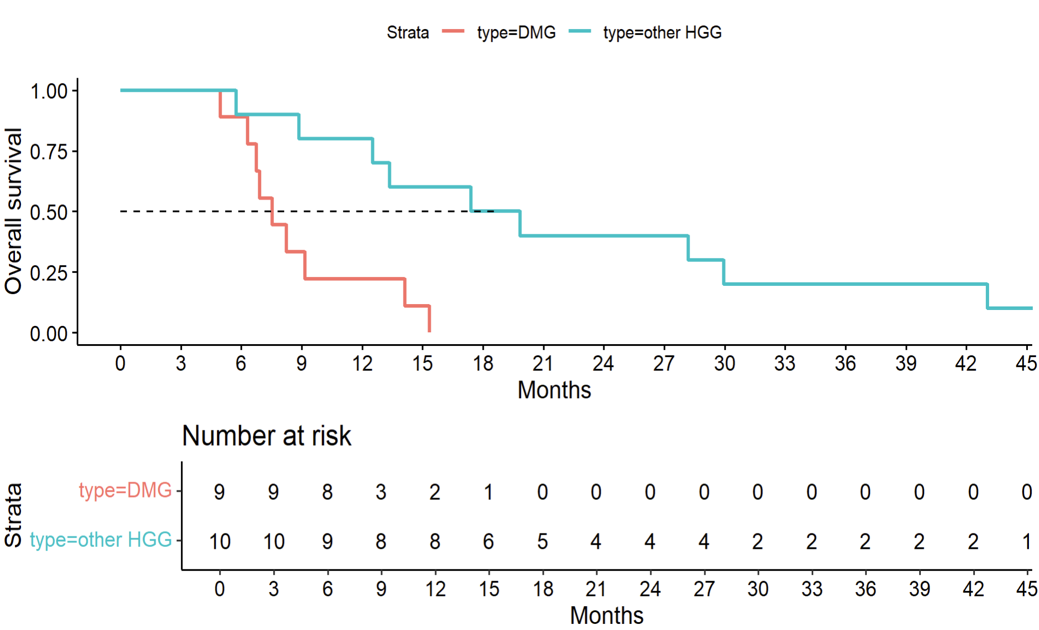

Supplementary material 2. Survival curves stratified by type of HGG (DMG versus other HGG).

|  | Adjusted HR (95% CI) | Adjusted p-value |
| --- | --- | --- |
| **Dox** |  |  |
| *No dox* | 1 | 0.55 |
| *Dox* | 0.74  (0.28-1.96) |  |
| **Type of HGG** |  |  |
| *DMG* | 1 | **0.01** |
| *Other HGG* | 0.21  (0.06-0.72) |  |
| Note: Dox: doxorubicin, HGG: high-grade glioma, DMG: diffuse midline glioma, HR: hazard ratio, CI: confidence interval | | |

Supplementary material 3. Results of multivariate Cox regression without the interaction term.

| Time  (months) | | 6 | 9 | 12 | 15 | 18 | 36 | 75.4  (End of follow up) |
| --- | --- | --- | --- | --- | --- | --- | --- | --- |
|  | **N** |  |  |  |  |  |  |  |
| **Overall** | 19 | 0.91  (0.78-1) | 0.57  (0.39-1) | 0.48  (0.30-0.75) | 0.33  (0.18-0.61) | 0.24  (0.11-0.51) | 0.10  (0.03-0.36) | 0.048  (0.01-0.32) |
| **Dox (I)** | 10 | 1 | 0.8  (0.59-1) | 0.7  (0.47-1) | 0.4  (0.18-0.85) | 0.3  (0.12-0.77) | 0.1  (0.02-0.64) | 0.1  (0.02-0.64) |
| *DMG* | 3 | 1 | 0.67  (0.3-1) | 0.33  (0.07-1) | 0 | - | - | - |
| *Other HGGs* | 7 | 1 | 0.86  (0.63-1) | 0.86  (0.63-1) | 0.57  (0.30-1) | 0.29  (0.09-0.92) | 0.14  (0.02-0.88) | 0.14  (0.02-0.88) |
| **No Dox** | 9 | 0.78  (0.55-1) | 0.33  (0.15-0.84) | 0.33  (0.15-0.84) | 0.33  (0.15-0.84) | 0.22  (0.07-0.75) | 0.11  (0.02-0.71) | 0 |
| *DMG* | 6 | 0.83  (0.58-1) | 0.17  (0.03-1) | 0.17  (0.03-1) | 0.17  (0.03-1) | 0 | - | - |
| *Other HGGs* | 3 | 0.67  (0.30-1) | 0.67  (0.30-1) | 0.67  (0.30-1) | 0.67  (0.30-1) | 0.67  (0.30-1) | 0.33  (0.07-1) | 0 |
| Note: Dox: doxorubicin, N: number of patients, DMG: diffuse midline glioma, HGG: high-grade glioma | | | | | | | | |

Supplementary material 4. Overall survival probabilities and survival probabilities stratified by Dox administration (I Dox dose or NO Dox).
